# *CTNNB1* (*β*-catenin) mutations in NSCLC: clinicogenomic characteristics, prognostic value and implications for therapy

**DOI:** 10.1101/2025.11.12.25338824

**Authors:** Moritz Glaser, Cornelia von Levetzow, Anna Rasokat, Darinka Prang, Lucia Nogova, Claudia Wömpner, Jaqueline Schmitz, Elisabeth Bitter, Inken Terjung, Anna Eisert, Rieke Fischer, Felix John, Sebastian Michels, Richard Riedel, Lea Ruge, Heather Scharpenseel, Wolfgang Schulte, Frank Beckers, Urte Sommerwerck, Udo Siebolts, Sabine Merkelbach-Bruse, Reinhard Buettner, Jürgen Wolf, Matthias Scheffler

## Abstract

Although mutations in *CTNNB1* have long been associated with cancer, their impact in patients with non-small cell lung cancer (NSCLC) is not well understood. Beyond a potential role in the acquired resistance setting of *EGFR*-mutant NSCLC, little is known about the clinical and molecular characteristics of NSCLC patients harboring these mutations. Here, we identify 302/15,688 (1.9%) NSCLC patients with *CTNNB1* mutations. These mutations frequently co-occur with *EGFR* and *KRAS* mutations and are associated with a favorable prognosis (mOS, 45.8 months overall; 19.7 months *KRASmut*). Patients benefit significantly from immune-checkpoint-blockade, but non-intuitively,PD-L1 TPS ≥50%(4/6 treated with monotherapy) survive the shortest. We show that patients with proposed *CTNNB1* resistance mutations to EGFR-directed therapy have significantly shorter mOS when treated with targeted therapy, compared to non-resistant *CTNNB1* mutations. This study highlights unique clinicogenomic features and the nuanced impact of *CTNNB1* mutations on therapeutic outcomes.

## 1. Introduction

Over the past decade, advances in molecular diagnostics and targeted therapies have transformed the treatment landscape for non-small cell lung cancer (NSCLC)^1,2^. Targeted therapies for specific driver aberrations such as ALK fusions have led to unprecedented progression-free survival (PFS) with a median PFS still not reached after 5 years of follow-up^3^. However, a comprehensive understanding of less common genetic alterations and their clinical implications remains incomplete^4^. Among these, mutations in the *CTNNB1* gene, which encodes *β*-catenin, represent a relatively long known but understudied subset of molecular aberrations in NSCLC^5–7^.

*CTNNB1* mutations are identified in approximately 2% of NSCLC cases, predominantly in adenocarcinomas. *β*-Catenin plays a central role in the Wnt/*β*-catenin signaling pathway, which is essential for cellular processes such as proliferation, differentiation and apoptosis^8^. Dysregulation of this pathway, often mediated by *CTNNB1* mutations, has been implicated in tumorigenesis in several cancers^9^. These mutations typically affect phosphorylation sites within the *β*-catenin protein, leading to its stabilization and nuclear accumulation, thereby promoting aberrant activation of Wnt target genes. Common mutations, including T41A, S37F, and S45F, disrupt *β*-catenin degradation and are thought to contribute to tumor initiation and progression^9^. Consequently, efforts have been made to target these mutations in *β*-catenin with small molecules^8^, either directly by inhibiting its transcriptional activity or indirectly by inhibiting associated enzymes^10,11^.

Despite the mechanistic insights gained from *CTNNB1* mutations in preclinical studies, their broader clinical relevance in NSCLC remains poorly defined. Research to date has primarily focused on their potential role in resistance to targeted therapies, particularly in the context of *EGFR*-mutant NSCLC. For example, early preclinical studies suggested that *CTNNB1* mutations may co-occur with *EGFR* mutations and lead to cross-talk between signaling pathways, potentially influencing drug sensitivity. One study researched the role of co-occurring mutations in *EGFRmut* advanced stage (III or IV) NSCLC with a genomic analysis of cell-free DNA samples^12^. According to the study, mutations in Wnt- and MAPK-pathway genes were significantly enriched in *EGFRmut* patients whereas gene fusions were significantly enriched in the *EGFRwt* cohort. Analyzing longitudinal exome sequencing of one patient’s tumor, the study finds that *CTNNB1 S37F* alongside an exon 19 deletion was already present in early-stage disease and not restricted to a specific lesion. A recent translational study scrutinized the role of different *CTNNB1* mutations in resistance to EGFR targeted therapy^13^. In the HCC827 cell line, a subset was identified that *“can cause resistance”* (including S37F) to first-generation TKI erlotinib, and a subset that does not cause resistance, apparently. *CTNNB1* T41A was most resistant and also showed *“sustained”* ERK and AKT signaling. An earlier translational study also identified *CTNNB1* T41A to confer significant resistance to targeted therapy in *EGFRmut* and *ALK* translocated cell lines (HCC827 and STE1)^14^.

Besides the vague evidence of contributing resistance to targeted therapy, no conclusion could be drawn regarding the prognostic impact of *CTNNB1* mutations in NSCLC due to small sample size^15,16^. The clinical and molecular landscape of *CTNNB1*-mutant NSCLC, including the types and frequencies of co-occurring driver or passenger mutations, remains largely unexplored. Co-mutations in genes such as *TP53, STK11* and *KRAS* are commonly observed in NSCLC and may interact with CTNNB1-driven tumorigenesis. In addition, there is limited data on how *CTNNB1* mutations influence disease stage, histologic characteristics or patient prognosis. These gaps highlight the need for comprehensive studies to better characterize the clinical profile of *CTNNB1*-mutant NSCLC patients.

Our recent work addresses these knowledge gaps by screening a large real-world cohort of NSCLC patients for *CTNNB1* mutations. Through this study, we aimed to describe the clinical features, molecular alterations, and potential prognostic implications associated with this subset of NSCLC patients. By focusing on the broader clinical and molecular context of *CTNNB1* mutations, we sought to provide novel insights into their significance in NSCLC and to lay the groundwork for further investigation of targeted therapeutic strategies.

## 2. Results

### 2.1 Patient Demographics

Of 15,688 NSCLC patients who underwent NGS sequencing for *CTNNB1* (**methods**) from January 2018 until June 2023, we identified 302 patients with *CTNNB1* mutation (1.9%, **Fig. 1A**) for which mOS was 45.8 months in stage IV patients (CI: 26.4-65.1, **Fig. 1B**).

**Figure 1:**
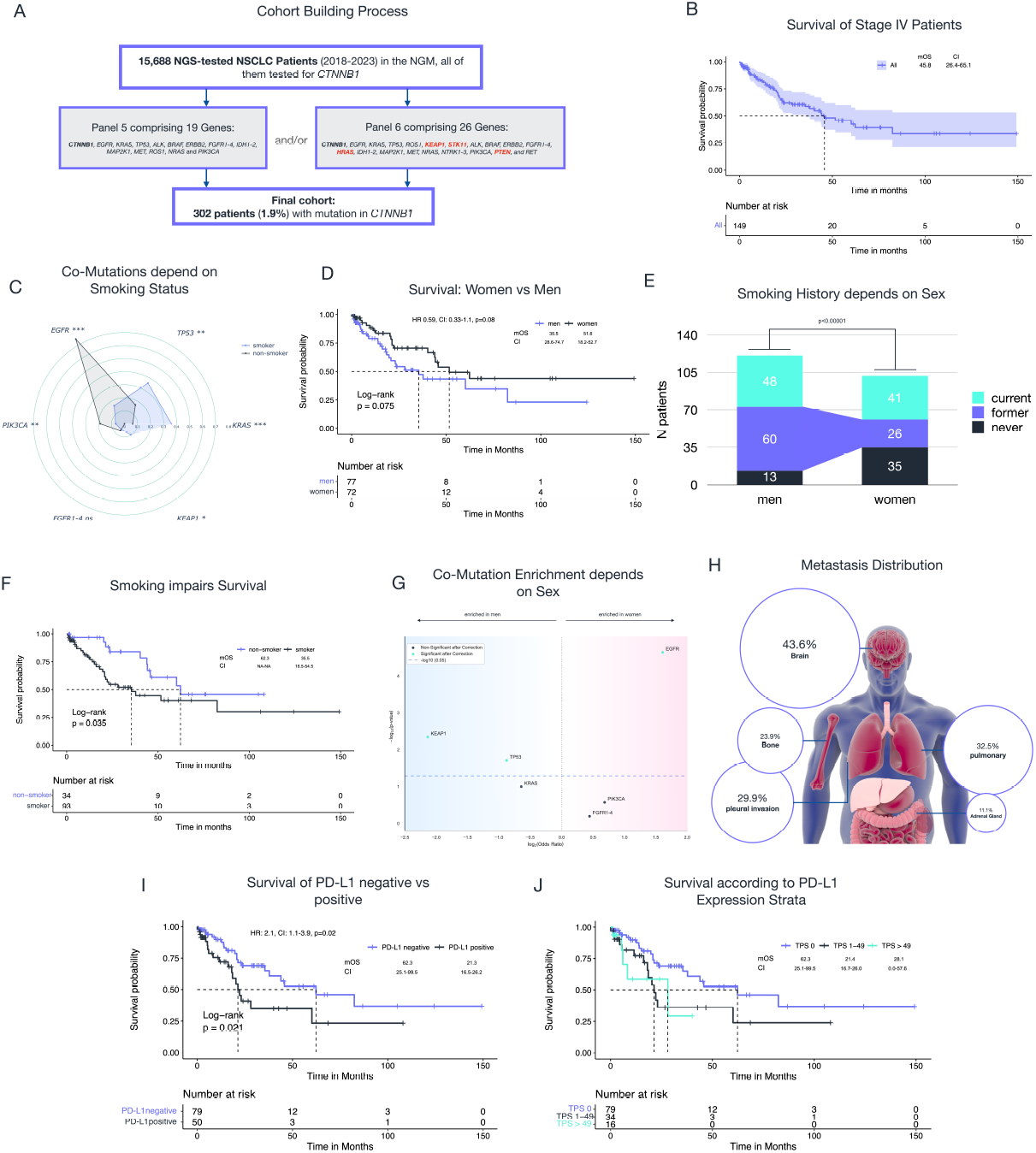
Clinical and molecular Characteristics of CTNNB1mut NSCLC patients and Survival. **A** shows the cohort building process of this study. **B** shows the survival of all stage IV patients with survival data available and above 1 months if not deceased (n=149/202). Median followup was 13.8 months (CI: 6.0-20.8). **C** is a radar chart contrasting the co-mutations of patients with smoking history with those patients without. All differences except for FGFR1-4 are significant with Benjamini-Hochberg correction (*** p<0.001, ** p<0.01, * p<0.05, ns not significant). While 95.5% of KRAS co-mutant (n=63/66), 88.7% of TP53 co-mutant (n=63/71) and 94.4% of BRAF (n=17/18) co-mutant patients were former smokers or current smokers (“smoking history”), only 57.1% of PIK3CAmut patients (12/21) and just 43.8% of EGFRmut patients (n=28/64) had a smoking history. **D** Survival comparison between men and women. **E** illustrates the difference in smoking status between women and men. Highly significant more men had a smoking history than women (p<0.00001). **F** survival comparison of stage IV patients according to binarized smoking history. **G** Volcano plot showing the correlation of different co-mutant genes with the sexes in all patients irrespective of stage and censor. P-values are corrected against multiple testing using the Benjamini-Hochberg procedure. **H** visualizes the metastasis distribution in a simplified patient model. Metastasis data was available for 117 of the 200 stage IV patients (58.5%) and indicates any presence of metastases since stage IV diagnosis. **I-J** survival according to binarized PD-L1 expression and PD-L1 TPS strata. Note that 60.1% of patients (n=153/251) did not express any PD-L1, 28.7% had a TPS of 1-49 (n=72/251) and 10.4% had a TPS greater than 49 (n=26/251). In 51 patients no expression data was available. Interestingly, when regarding deceased stage IV patients only, the highest TPS among men was 100, while 30 among women. But regarding all patients, the difference between the sexes was not significant (p=0.99).

Clinical characteristics are summarized in **Table 1**: 54.3% of patients were males (n=164/302) who survived shorter than women (35.5 [CI: 28.6-74.7] vs 51.6 months [CI: 18.2-52.7], p=0.075) (**Fig. 1D**). More men had a smoking history than women (p<0.00001): 34.3% of women were never smokers, while only 10.7% of men have never smoked (**Fig. 1E**). Smoking shortened survival significantly (mOS 35.5 vs 62.3 months, CI: 16.5-54.5 vs NA-NA, p=0.035; HR 2.2, CI: 1.0-4.5, **Fig. 1F**). Smokers had significantly more *KEAP1, TP53* and *KRAS* mutations (**section 2.3**.) and men had significantly more *KEAP1* and *TP53* mutations (*KRAS* not significant (ns)), while women had more *EGFR* mutations and non-smoker had significantly more *EGFR* and *PIK3CA* co-mutations (**Fig. 1C, 1G**).

The median age at first diagnosis was 67 years (range: 34-90) and there was no significant difference between the sexes (p=0.14). The most common histological type was adenocarcinoma (AD), with 257 patients (91.1%) diagnosed, followed by squamous cell carcinoma (SqCC) (6.0%, n=17).

Most patients with stage IV (68.3%, n=170/249) had brain metastases at first diagnosis (43.6%; n=51/117, **Fig. 1H**), whereof 31.4% had an *EGFR* mutation (versus 28.5% in the overall cohort). *KEAP1* mutations were significantly (p=0.03) enriched in patients with brain metastases: 7 of 8 (87.5%) patients with *KEAP1* mutations had brain metastases versus 51 of 117 patients (43.6%) with any co-mutation.

### 2.2. Molecular coordinates and survival

Most patients (n=153/251, 60.1%) did not express any PD-L1 and 28.7% had a Tumor Proportion Score (TPS) of 1-49% (n=72/251). Patients with PD-L1 negative tumors survived significantly longer than PD-L1 positive: patients with expression had an mOS of 21.3 months (CI: 16.5-26.2) versus 62.3 (CI: 25.1-99.5) months without expression (**Fig. 1I** and **1J** for different strata, p=0.02, HR: 2.1, CI: 1.1-3.9). *TP53* mutations (**section 2.3**.) were significantly enriched in patients with PD-L1 expression (log2(Odds Ratio): 1.5, p<0.0005), while *EGFR* mutations were slightly enriched in patients without (log2(Odds Ratio): 0.6, p=0.6) and the other mutations did not show any association. Thus, the favorable outcome of patients without PD-1 expression is likely driven by *EGFR* mutations and EGFR-targeted therapy (**section 2.7**).

**Figure 2A** and **2C** show a detailed picture of mutation type and position: of the 322 *CTNNB1* mutations in total (multiple mutations in some patients), most were missense and predicted oncogenic. Mutations at position S37 were most common (42.2%; n=137/322) followed by S33 (15.2%; n=49/322) and S45 (14.0%; n=45/322, **Fig. 2A**). The most frequent *CTNNB1* mutation was S37F (n=77), followed by S37C (n=51), S45P (n=23) and S33C (n=22). Mutations at position 37 were most common irrespective of the co-mutation (**Fig. 3A**) and the PD-L1 TPS (**S1** (Supplementary Figure 1)) and OS did not show a clear correlation with the position of the mutant *CTNNB1* residue (**Fig. 3B**).

**Figure 2:**
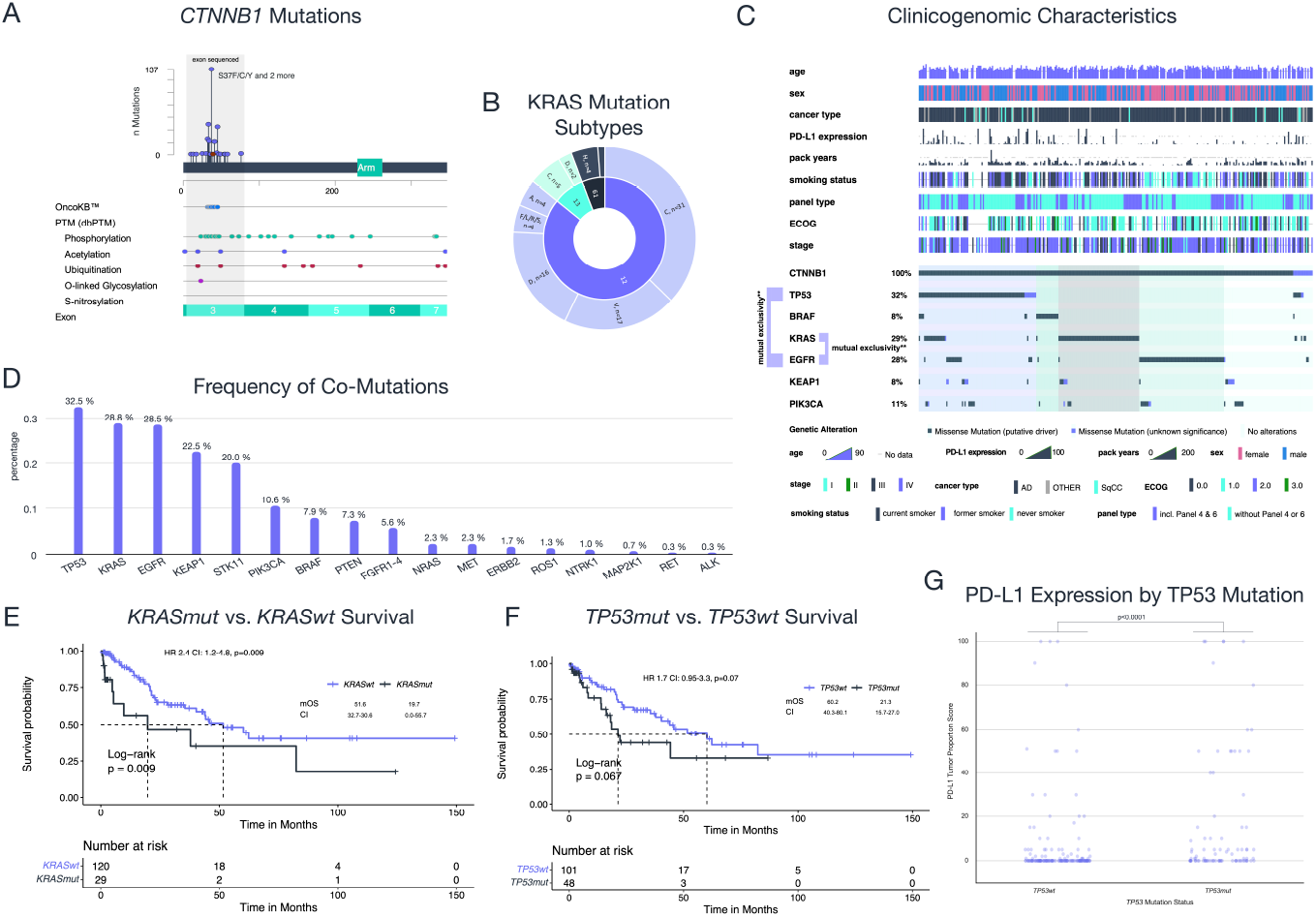
Molecular Characteristics and Survival I. **A** shows a lollipop chart of the CTNNB1 mutations in this cohort, displaying their position and frequency in the gene. Overall, almost all mutations (98.1%) were missense mutations - only 4 in-frame deletions and 2 truncating mutations have been found. According to OncoKB, 79.9% of mutations were considered “likely oncogenic” (n=257/322), 13.4% of mutations classified “oncogenic” (n=43/322), 22 mutations were unknown. **B** shows the three most commonly mutated KRAS residue positions and their respective substitute when mutated (only positions with more than one mutation are shown). **C** is an Oncoprint showing clinicogenomic data of the cohort. Both mutual exclusivities are p<0.001 and significant with Bonferroni correction. **D** shows the frequency of mutations among the different co-mutated genes in descending order. 22.5% had KEAP1 mutations (23 of 102 patients tested) and 20.0% of patients had STK11 mutations (18 of 90 tested). **E-F** compare the survival of patients with or without KRAS or TP53 co-mutation, respectively. **G** illustrates the significant difference in PD-L1 TPS between TP53mut and TP53wt patients. Median TPS in TP53wt was 0, and 1 in TP53mut patients.

**Figure 3:**
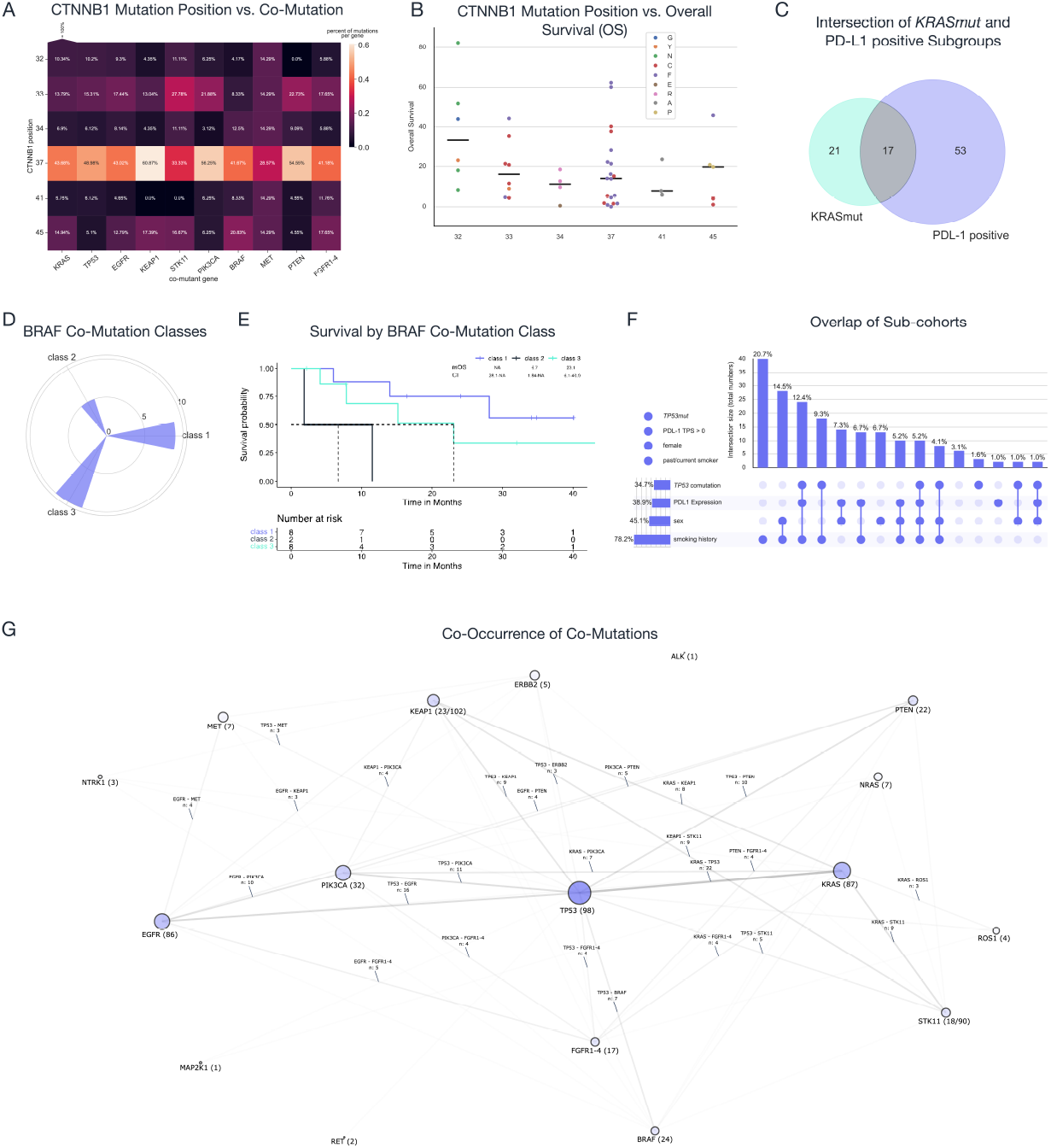
Molecular Characteristics and Survival II. **A** Heatmap showing with which mutant CTNNB1 position each co-mutant gene occurs most frequently. All columns are normalized by the absolute number of co-mutations for the respective gene and sum up to 100% column-wise. **B** shows the variation in overall survival across the different CTNNB1 residues affected by mutation with each datapoint being color-coded by its respective substituting amino acid. Only deceased patients are included in this analysis. The survival difference of patients with T41 and non-T41 mutations was far from significant (p=0.87). **C** Venn diagram showing the overlap between the two groups with significant survival differences (KRAS co-mutated and PD-L1 positive). **D** radial bar chart showing the absolute frequencies of the different BRAF mutations classes. Of the patients with class 3 mutations, 3 had a TP53 mutation, but KRAS and EGFR occurred only once respectively. **E** survival according to BRAF co-mutation. **F** UpSet plot showing the intersections between the different sub-cohorts. Light-colored dots represent the opposite of the property specified in the legend. G Graph network showing which co-mutations occur how often with each other. Interestingly, TP53 co-mutations are mutually exclusive with EGFR mutations (p<0.01), as are KRAS and EGFR (p<0.001). The absolute numbers of patients with co-mutation are shown in brackets, but STK11 and KEAP1 were not tested in all patients. Size and opacity of nodes and links are optimized for better readability and do not reflect the true absolute numbers and the spring layout algorithm was used (cf. methods).

### 2.3 Co-mutations and sub-cohort overlaps

Co-occurring mutations as detected by the panels (**Fig. 1A**) are shown in **figure 2D**: 6.3% of patients had a mutation in *CTNNB1* (n=19/302) as the only mutation, 32.5% of patients had a *TP53* mutation (n=98) and 28.8% of patients had *KRAS* mutations (n=87, therefrom 31 with G12C, **Fig. 2B**). *TP53* mutations (mOS 21.3 vs 60.2, CI: 15.7-27.0 vs 11.9-63.7, p=0.11; HR 0.6, CI: 0.3-1.1) and *KRAS* mutations (mOS 19.7 vs 51.6, CI: 0.0-55.7 vs 32.7-30.6; p=0.009; HR 2.4 CI: 1.2-4.8; **Fig. 2E-F**) shortened survival. There was no notable survival difference between KRAS G12D and G12C mutant patients (p=0.93) which were the most common KRAS subtypes together with G12V (**Fig. 2B**).

To investigate interactions, we calculated the respective intersections of PD-L1 expression and the presence of KRAS mutations (**Fig. 3C)**, showing that 17 of 70 PD-L1 positive patients had a *KRAS* mutation (24.3%), and 17 of 38 *KRASmut* patients expressed PD-L1 (44.7%) revealing minor intersections.

Besides, 28.5% of patients had an *EGFR* mutation (n=86, **Fig. 2D)**. Of the *PIK3CA* mutations (10.6%), 75.0% were in exon 9 (n=24/32), the remaining in exon 20 (**S4**). Of all *MET* mutations (n=7), four were point mutations (none annotated), deletions, duplications and high-level amplifications (GCN 6.2) occurred once. *BRAF* mutation classes are shown in **figure 3D** and the survival in **figure 3E**. 34.8% of patients (n=105/302) had RAS/RAF pathway activating mutations only and 12.9 in the PI3K path way exclusively (**supplement**). Patients with RAS pathway alteration and additional PI3K pathway alteration (n=6/48) survived shorter (mOS 7.3 vs. 23.1 months, CI: 5.5-40.6 vs 0.0-20.7, p=0.2) than patients with a RAS pathway alteration only.

Overall, this leaves 62.9% of patients without mutations in a targetable gene (n=190/302 in *EGFR, MET* and *BRAF*) and 26.5% without additional oncogenic mutation (n=80/302). We reconstruct which co-mutations often occur in a graph (**Fig. 3G**) showing that *TP53, KRAS* and *EGFR* rarely occured together, especially in the combination *KRAS*-*EGFR* (n=2, p<0.001). But the co-occurrence of *TP53* and *EGFR* mutation was unlikely (p=0.01) as well. In addition, we elucidate the intersections of the different sub-cohorts of our patients (**Fig. 3F**): smoking male patients were the most prevalent subcohort by a considerable margin (20.7%).

Besides, a multivariate analysis of univariately significant clinical characteristics, co-mutations and PD-L1 expression is provided in **figure 4C**: indeed, PD-L1 positivity and *KRAS*mut were multivariate significant, too.

**Figure 4:**
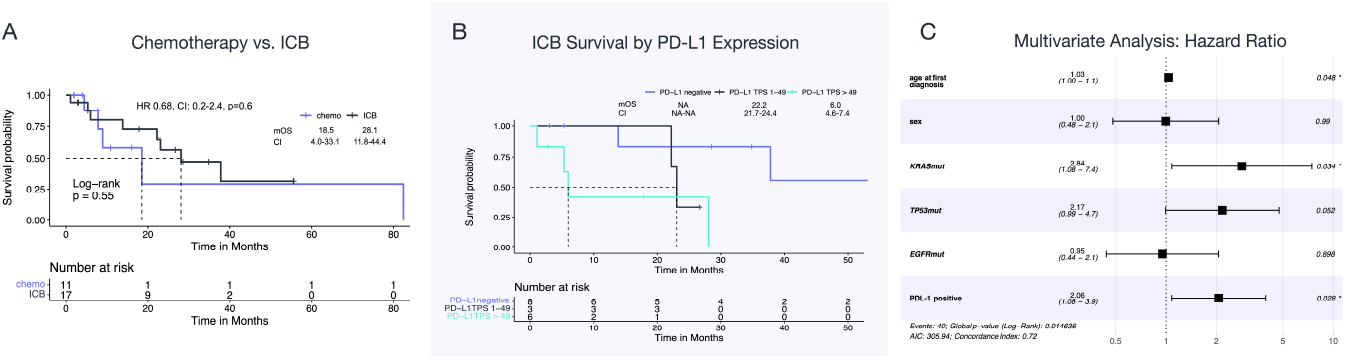
Influence of Therapy on Overall Survival. **A** shows the survival according to non-targeted treatment (ICB vs. chemotherapy). **B** shows the survival of patients treated with ICB by the PD-L1 expression stratum. **C** shows a forest plot of a multivariate analysis (Cox regression) according to main characteristics relevant for survival.

### 2.5 Benefit from ICB in patients without clinically actionable drivers

*BRAF-*STKI naive patients with *EGFRwt* (incl. *KRASmut*) receiving ICB (n=17) during treatment survived 10 months longer than patients only treated with chemotherapy (n=11) and/or angiogenesis inhibition (**Fig. 4A**, mOS 28.1 vs 18.5 months, CI: 11.8-44.4 vs 4.0-33.1, p=0.55; HR 0.7, CI: 0.2-2.4).

ICB benefit significantly depended on PD-L1 expression in a non-intuitive manner: patients with PD-L1 positive tumors had an mOS of 22.2 months while patients with negative tumors did not reach mOS (n=8, CI: 2.5-41.9 vs NA-NA, p=0.017; HR: 9.5 CI: 1.1-84.1). Patients with TPS ≥ 50, survived particularly shot (mOS 6.0 months,CI: 4.6-7.4, n=6) followed by TPS 1-49 (mOS 23.1, CI: 21.7-24.4, n=3, **Fig. 4B**). However, sample sizes were too small to draw robust conclusions.

### 2.6. Role of CTNNB1 mutations in EGFR TKI resistance

Among *EGFR* mutations (28.5%) exon 19 deletions were most common (41.4%; n=48/116), wherefrom 28 were E746_A750 deletions (**Fig. 5A**). 16 patients had an L858R mutation and 13 acquired T790M mutations after first- or second generation EGFR inhibition. Patients with *EGFR* mutations had prolonged survival (mOS 51.6 vs 37.8, CI: 30.2-73.0 vs 11.9-63.7, p=0.11; HR 0.6, CI: 0.3-1.1, **Fig. 5C**), and the type of mutation (E746_A750del or L858R) did not make a significant difference to survival (p=0.15, p=0.17 for L858R vs non L858R and E746_A750del vs non-E746_A750del). Patients with exon 19 deletion survived significantly longer than patients without (p=0.0034, n=36/60).

**Figure 5:**
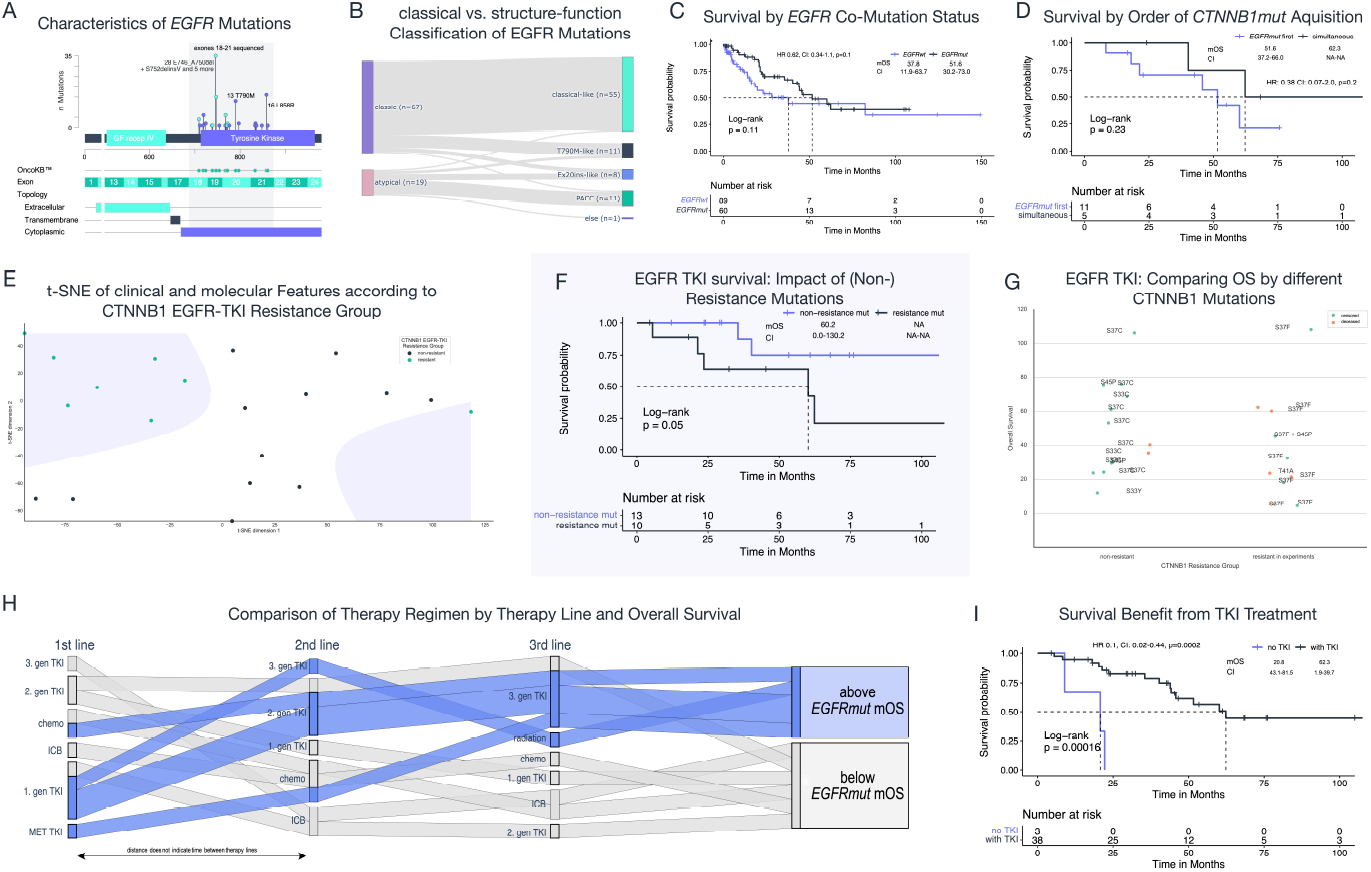
EGFR Analysis: Mutation characteristics, Survival Differences for Resistance Mutations and Therapy. **A** is a lollipop chart of all EGFR mutations in the cohort (28.5%, n=86/302). Exons 2-13 are truncated as they were not included in the panel. 66.4% of mutations (n=22) were “oncogenic”, the impact of the other 9 mutations is “unknown”. 16 patients had an L858R mutation and 13 had a T790M mutation. **B** shows the classification of EGFR mutations according to the traditional binary scheme and the structure-function based classification proposed by Robichaux et al. **C-D** survival of patients with and without EGFR co-mutation and whether the EGFR mutation occurred before the CTNNB1 mutation or simultaneously. **E** visualizes a t-distributed stochastic neighbor embedding (t-SNE) of the most important clinical and molecular characteristics of 20 patients such as survival (only deceased patients included), PD-L1 expression, age, ECOG performance state, smoking status, frequent co-mutations etc. (details in methods). The contours outline the decision boundaries of a polynomial regression model of second degree. **F** survival difference of EGFRmut patients treated with an EGFR TKI comparing patients with pre-clinically non-EGFR TKI resistant vs. resistant mutations finding a significant difference. **G** shows the difference in OS of EGFRmut patients treated with a TKI, grouped by their type of CTNNB1 mutation (shown to confer resistance experimentally vs non-resistant) and color-coded by their censor. **H** shows the treatment regimen and corresponding outcome for stage IV EGFRmut patients with 3 lines of therapy data available (n=11). **I** shows the survival difference between EGFRmut patients treated with vs without TKI.

**Figure 5B** classifies *EGFR* mutations according to the binary (“classical” versus “atypical”^17^) and the structure-function classification, finding that the vast majority of mutations were “classical” respectively “classical-like” mutations^18^. Four patients with “exon 20 insertion like” survived the shortest (mOS 22.2, CI: 7.7-36.8, **supplement** for other classes).

Fathoming why some *CTNNB1* mutations cause EGFR targeted therapy resistance preclinically and some not, we found that AlphaMissense pathogenicity scores for predicted *CTNNB1* resistance mutations are lower on average (0.92 vs. 0.98), though the difference is miniscule (p=0.8, **S2**) and the underlying structure predictions are low in confidence (**S3, methods**). Likewise, the (significant) predicted increase in protein stability by the CUPSAT method of potential resistance mutations is uncertain, too (**supplement**).

Three of four resistance-predicted mutations affect the same position as their non-resistant-predicted counterparts. Despite non-significance, the PD-L1 TPS differed considerably between non-resistant and resistant groups (p=0.09, median both 0, mean: 8.1 vs 13.6). We use dimensionality reduction and polynomial regression to inquire about the overall cohort difference between these two mutational groups, finding the predicted resistance mutations to have a distinct embedding (**Fig. 5E**).

### 2.7. Impact of CTNNB1 mutations on survival in EGFR-mutated and EGFR TKI-treated patients

A third of patients acquired the *CTNNB1* mutation after the *EGFR* mutation (n=7/21), but in 61.9% of patients both mutations occured simultaneously (n = 13/21, **Fig. 5D**). The mOS of patients with simultaneous mutation was 11 months shorter (51.6 months, CI: 37.2-66.0) compared to patients with *EGFR* mutation preceding the *CTNNB1* mutation (mOS 62.3, CI: NA-NA, p=0.23; HR: 0.4, CI: 0.1-2.0). All of the patients with preceding *EGFR* mutation were treated with a TKI and received osimertinib over the course of the treatment, two of them in the first line. Two patients also received erlotinib. 12 of 13 patients with simultaneous *EGFR* and *CTNNB1* mutation were treated with TKI.

As expected, *EGFRmut* patients treated with TKI (n=38/43) survived significantly longer than those treated without (**Fig. 5I**, mOS 62.3 vs 20.8, CI: 43.1-81.5 vs 1.9-39.7, p<0.001, HR 0.1, CI: 0.0-0.4).

We compared the survival of TKI-treated *EGFRmut* patients with a *CTNNB1* mutation belonging to the predicted resistance subset ((T41A, S37F, S45C and D32H), n=10)^13^ versus mutations assumed not to confer resistance ((D32Y, S33C, S33Y, S45P, and S37C), n=13, **Fig. 5F-G**)^13^. Patients in the latter group survived significantly longer (mOS 60.2 vs NA, CI: 0.0-130.2 vs NA-NA, p=0.05). 4 of 10 of patients in the group with predicted resistance mutation did express PD-L1, while 2 of 11 patients with non-resistance mutation expressed PD-L1.

## 3. Discussion

The aim of this study was to evaluate the clinicogenomic characteristics of this subcohort including survival and treatment dependencies, its suitability for targeted therapy and its role in EGFR TKI resistance from a clinical perspective.

Regarding the first objective, this work represents the most comprehensive characterization of *CTNNB1mut* NSCLC to date, analyzing 302 of 15,688 patients screened. We were able to define this subset of NSCLC: it is an older, slightly more male, smoking subset with predominantly adenocarcinomas, almost 50% brain metastases, homogeneous *CTNNB1* mutations independent of co-mutations and generally low PD-L1 expression dependent on *TP53* co-mutation.

However, the most striking finding of this study is the very complex heterogeneity of the cohort, both clinically and molecularly. The mutations occur in both heavy-smokers and never-smokers, which is reflected in an almost contradictory set of oncogenic drivers: On the one hand, they co-occur, as previously described^12^, in patients with *EGFR* mutations, on the other hand, in patients with *KRAS* mutations; both practically equally frequent. It is therefore necessary to analyze these patient subgroups separately, with focus on targeted therapy in the EGFR group and on ICB in the KRAS group plus the remainder.

Nevertheless, there are some aspects shared by this heterogeneous cohort: The mOS in stage IV independent of co-mutation is impressive, especially considering the fact that *EGFR* and *KRAS* mutations are equally common. mOS was above expectations in both the EGFR cohort (62.3 months) and the KRAS cohort (19.7 months). We observed a benefit from ICB in the entire cohort, but without correlation to PD-L1 expression. Although our data are based on small sample sizes that do not allow for strong conclusions, they do not suggest a lack of efficacy of ICB in our cohort. However, we found non-intuitive results regarding the association of ICB with PD-L1 expression.

Our analysis of the impact of mutations on EGFR targeting was ambivalent: we showed clearly that *CTNNB1* mutations are not exclusively acquired during treatment, but are often present in the first clone. Together with the finding that *CTNNB1* mutation acquisition after *EGFR* mutation only moderately shortens survival, this argues against a role for these mutations in promoting resistance. Nonetheless, a subset of mutations which were preclinically described as promoting resistance in patients treated with erlotinib shortens survival of TKI treated patients significantly and elevates prognostic PD-L1 expression^13,14^.

In addition to the small sample sizes of our subgroups due to the low frequency of these mutations, our study has several limitations especially regarding the assessment of treatment outcomes. It is a retrospective analysis of a real-world cohort, where data collection is not comparable to that of prospective clinical trials. We were unable to collect treatment details such as radiographic response or PFS for specific therapies and were therefore limited in our analysis to OS.

In conclusion, this study adds evidence about *CTNNB1* mutations not only as a potential mode of resistance in *EGFR*-mutated NSCLC but also of co-occurring *KRAS* mutations.

## 4. Methods

### 4.1. Patients

Formalin-fixed paraffin-embedded tumor-tissue samples were analyzed by the Institute of Pathology of the University Hospital of Cologne. All patients provided written informed consent and patient data was hand-curated by an experienced oncologist. The study was reviewed by the institutional ethics committee.

### 4.2. Molecular diagnostics

Exon 3 of the *CTNNB1* proto-oncogene was sequenced in two different panels (LUN 5 & 6, **Figure 1**) and exons 18-21 of *EGFR* were sequenced. Only results with a coverage >200 were interpreted and a 5% cutoff for variant cells was utilized. The different NGS panels and genes covered are shown in **Figure 1** (further information in **supplementary table 1**). Programmed-cell-death ligand-1 (PD-L1) expression was assessed in the Tumor Proportion Score (TPS) format.

### 4.3. Clinical parameters

Age, sex, stage, smoking status, smoking quantity (pack-years), OS, therapy data and outcome were assessed when data was available. Thus percentages and fractions are always adapted to the number of patients with data available. The median followup was 15.7 months using the “reverse Kaplan Meier method”. Patients with less than <100 cigarettes classified as never-smokers, smoking cessation longer than two years was criterion for ‘former smokers’. Eastern Cooperative Oncology Group’s (ECOG) performance status assessed patients’ condition. Disease and medical history was assessed in accordance with the treating physicians by reviewing patient records. Tumor stage at first diagnosis indicated by the Union for International Cancer Control (UICC) TNM classification, 8^th^ edition, metastasis and response to treatment were assessed pursuant to local standards including Response Evaluation Criteria in Solid Tumors (RECIST) 1.1. Censored patients (i.e. without an event) with an OS below one month were excluded from the survival analysis.

### 4.4. Evaluation of detected variants

The impact of *CTNNB1, EGFR* and *MET* mutations was assessed using OncoKB knowledge base and divided into either oncogenic and non-oncogenic^26,27^. Patients with multiple *CTNNB1* mutations were excluded from analyses where the exact mutation was of interest. Mutation impact was evaluated using AlphaMissense predicting the pathogenicity of a mutation from evolutionary data. Its scores were accessed from the original publication^22^. The CUPSAT method was used for protein stability prediction upon point mutation (**supplement**)^23^. *BRAF* mutations were grouped into classes according to Owsley et al.^24^. *KRAS* mutations were not counted as targetable alterations.

### 4.5. Statistical Analyses

Qualitative and quantitative variables are displayed by count and percentage. Survival was calculated using Kaplan-Meier methods. The “Reverse Kaplan Meier” method was used to assess median follow-up (mFU) and was 23.7 months (CI: 13.8-33.4). Log-rank tests were performed for survival analysis, Chi-squared tests for categorical variables (e.g. smoking difference among the sexes), but Fisher’s Exact test was used when the lowest single frequency below 5. Man-Whitney test was used for the comparison of quantitative variables and Cox-proportional hazard model for hazard ratios (HRs). As in none of the analyses p-values differed between the HRs from the Cox regression and log rank Kaplan-Meier test, only the logrank p-value is indicated in the results. Confidence Intervals (CIs) were always set to 95%. P values below 0.05 were considered significant. Benjamini-Hochberg correction against multiple testing was used for the sex/brain metastasis/smoking-co-mutation association test (False-Discovery Threshold 0.05) and Bonferroni correction was used for the mutual exclusivity tests^25^. In the former association test KRAS, TP53, EGFR, PIK3CA, FGFR1-4 and KEAP1 were tested as they were of particular interest in the context of our analyses. Apart from that, no further corrections for multiple testing were conducted. Patients with multiple CTNNB1 mutations were discarded from analyses in which they would have been relevant due to ambiguity. The graph network representation of co-mutations uses the force directed spring layout algorithm with 38 iterations (spring constant = 3.5). For the t-distributed stochastic neighbor embedding (t-SNE), perplexity was set to 5 and dimension were reduced to 2 for visualization and a polynomial regression model with 2 degrees was used. For data analysis and visualization Affinity Designer (version 1.10.8), pandas (version 2.1.4.), seaborn (version 0.12.2.), Matplotlib (version 3.8.0.), Plotly (version 5.19.0), Numpy (version 1.24.3.), Scipy (version 1.11.4.), SciKit-Learn (version 1.2.2.), NetworkX (version 3.1.), RStudio (version 2023.12.1., RStudio Inc., Massachusetts, USA) and SPSS (version 28, IBM, Armonk, New York, USA) were used.

### 4.5. Code and Data availability

Alongside the supplementary information and figures we share the raw and anonymized patient dataset underlying this study. Moreover we share the R and python code for select analyses and figures. Code and dataset are deposited in the following github repository (github.com/moritzgls/CTNNB1-Paper-M-Glaser-et-al.).

## Data Availability

All data and code are available online at github.com/moritzgls/CTNNB1-Paper-M-Glaser-et-al.

## References

1. Scheffler, M., Michels, S. & Nogova, L. [Targeted treatment of non-small cell lung cancer]. Inn. Med. Heidelb. Ger. 63, 700–708 (2022).

2. Siegel, R. L., Miller, K. D., Wagle, N. S. & Jemal, A. Cancer statistics, 2023. CA. Cancer J. Clin. 73, 17–48 (2023).

3. Solomon, B. J. et al. Lorlatinib Versus Crizotinib in Patients With Advanced ALK -Positive Non–Small Cell Lung Cancer: 5-Year Outcomes From the Phase III CROWN Study. J. Clin. Oncol. 42, 3400–3409 (2024).

4. Skoulidis, F. & Heymach, J. V. Co-occurring genomic alterations in non-small-cell lung cancer biology and therapy. Nat. Rev. Cancer 19, 495–509 (2019).

5. The Clinical Lung Cancer Genome Project (CLCGP) and Network Genomic Medicine (NGM),. A Genomics-Based Classification of Human Lung Tumors. Sci. Transl. Med. 5, (2013).

6. Kraus, C. et al. Localization of the Human β-Catenin Gene (CTNNB1) to 3p21: A Region Implicated in Tumor Development. Genomics 23, 272–274 (1994).

7. Nusse, R., Van Ooyen, A., Cox, D., Fung, Y. K. T. & Varmus, H. Mode of proviral activation of a putative mammary oncogene (int-1) on mouse chromosome 15. Nature 307, 131–136 (1984).

8. Anastas, J. N. & Moon, R. T. WNT signalling pathways as therapeutic targets in cancer. Nat. Rev. Cancer 13, 11–26 (2013).

9. Sunaga, N. et al. Constitutive activation of the Wnt signaling pathway by CTNNB1 (beta-catenin) mutations in a subset of human lung adenocarcinoma. Genes. Chromosomes Cancer 30, 316–321 (2001).

10. Moroney, M. R. et al. Inhibiting Wnt/beta-catenin in CTNNB1 -mutated endometrial cancer. Mol. Carcinog. 60, 511–523 (2021).

11. Zaman, G. J. R. et al. TTK Inhibitors as a Targeted Therapy for CTNNB1 ( β -catenin) Mutant Cancers. Mol. Cancer Ther. 16, 2609–2617 (2017).

12. Blakely, C. M. et al. Evolution and clinical impact of co-occurring genetic alterations in advanced-stage EGFR-mutant lung cancers. Nat. Genet. 49, 1693–1704 (2017).

13. Majumder, A. et al. Abstract B087: CTNNB1 mutations can mediate resistance to EGFR targeted therapies in Non-Small Cell Lung Cancer. Mol. Cancer Ther. 22, B087– B087 (2023).

14. Majumder, A., Meyer, B. S., Hicks, J. K., Boyle, T. A. & Haura, E. B. Abstract 1101: CTNNB1 mutation can mediate resistance to EGFR, ALK and KRAS targeted therapies. Cancer Res. 82, 1101–1101 (2022).

15. Zhou, C., Li, W., Shao, J., Zhao, J. & Chen, C. Analysis of the Clinicopathologic Characteristics of Lung Adenocarcinoma With CTNNB1 Mutation. Front. Genet. 10, 1367 (2020).

16. Ros Montana, F. J. et al. WNT pathway mutations (APC/CTNNB1) and immune checkpoint inhibitors (ICI) response in metastatic non-small cell lung cancer (NSCLC) patients. Ann. Oncol. 30, v799 (2019).

17. Janning, M. et al. Treatment outcome of atypical EGFR mutations in the German National Network Genomic Medicine Lung Cancer (nNGM). Ann. Oncol. 33, 602–615 (2022).

18. Robichaux, J. P. et al. Structure-based classification predicts drug response in EGFR-mutant NSCLC. Nature 597, 732–737 (2021).

19. Jeanson, A. et al. Efficacy of Immune Checkpoint Inhibitors in KRAS-Mutant Non-Small Cell Lung Cancer (NSCLC). J. Thorac. Oncol. 14, 1095–1101 (2019).

20. Scheffler, M. et al. K-ras Mutation Subtypes in NSCLC and Associated Co-occuring Mutations in Other Oncogenic Pathways. J. Thorac. Oncol. 14, 606–616 (2019).

21. Jaiyesimi, I. A. et al. Therapy for Stage IV Non–Small Cell Lung Cancer Without Driver Alterations: ASCO Living Guideline, Version 2023.3. J. Clin. Oncol. 42, e23–e43 (2024).

22. Cheng, J. et al. Accurate proteome-wide missense variant effect prediction with AlphaMissense. Science 381, (2023).

23. Parthiban, V., Gromiha, M. M. & Schomburg, D. CUPSAT: prediction of protein stability upon point mutations. Nucleic Acids Res. 34, W239–W242 (2006).

24. Owsley, J. et al. Prevalence of class I-III BRAF mutations among 114,662 cancer patients in a large genomic database. Exp. Biol. Med. Maywood NJ 246, 31–39 (2021).

25. Benjamini, Y. & Hochberg, Y. Controlling the False Discovery Rate: A Practical and Powerful Approach to Multiple Testing. J. R. Stat. Soc. Ser. B Stat. Methodol. 57, 289–300 (1995).

26. Chakravarty, D. et al. OncoKB: A Precision Oncology Knowledge Base. JCO Precis. Oncol. 1–16 (2017) doi:10.1200/PO.17.00011.

27. Suehnholz, S. P. et al. Quantifying the Expanding Landscape of Clinical Actionability for Patients with Cancer. Cancer Discov. 14, 49–65 (2024).

