## Supplementary Material for "*CTNNB1* (*β*-catenin) mutations in NSCLC: clinicogenomic characteristics, prognostic value and implications for therapy"

### Supplementary Tables and Information

#### Table of content

**Information**

Impact of *MET* co-mutation on EGFR targeted therapy

Analysis of RAS/RAF vs PI3K pathway alterations

Survival according the structure-function *EGFR* mutation class

BRAF targeted therapy

Analysis of factors driving short survival in *EGFRmut* patients

Analyzing stability changes

**Figures**

S1

S2

S3

S4

S5

**Table**

NGS LUN Panels

### Information

#### Impact of *MET* co-mutation on EGFR targeted therapy

We also analyzed the impact of *MET* co-mutations on the survival of *EGFR* co-mutant patients treated with TKI. Interestingly, the two *METmut* patients did not experience an event (OS censored: 42.7 and 68.2 months), while the mOS of the remaining *EGFRmut* + *METwt* co-mutant patient was 60.2 months (CI: 44.2-NA , p=0.28). Therapy data was only available for one patient with a high-level *MET* amplification who was first treated with a MET inhibitor, later on with a third-gen EGFR inhibitor and responded to both.

#### Analysis of RAS/RAF vs PI3K pathway alterations

Analyzing the pathways affected by the co-mutations we found that besides the Wnt-pathway, 105 patients had co-mutations exclusively activating the RAS/RAF/MEK/ERK pathway, 39 patients with co-mutations exclusively activating the PI3K pathway and 10 patients had mutations stimulating both pathways. Note that RTKs and other genes were excluded. Patients with *KRAS* co-mutation mainly had RAS pathway alterations only (n=80/87), only 7 patients had a PI3K pathway alteration simultaneously. *BRAF* mutations behaved likewise. Similarly, of the patients with *PIK3CA* mutation only 9 of 32 had a RAS pathway alteration simultaneously. Patients with *TP53* co-mutations had a more diverse distribution with 26 RAS-only, 16 PI3K-only and 5 RAS + PI3K alterations.
For this analysis, *KRAS*, *MAP2K1*, *BRAF* and *NRAS* mutations count as RAS/RAF/MEK/ERK pathway mutations while *PIK3CA* and *PTEN* mutations are classified as PI3K pathway mutations. We excluded Receptor Tyrosine Kinases and other intracellular proteins deliberately for the clarity of the analyses.

##### Survival according to structure-function *EGFR* mutation class

We classified the *EGFR* mutations according to Robichaux et al. and compared their survival finding that patients with “exon 20 insertion like” mutations survived the shortest (mOS 22.2 months, CI: 7.7-36.8, n=4). mOS was 51.6 months in patients with “T790M like” mutation (CI: 43.4-59.9, n=8), 62.3 months with “classical like” (CI: NA-NA, n=38) and not reached in patients with “P-loop and alpha helix compressing (PACC)” mutation (CI: NA-NA, n=7).

**BRAF targeted therapy**

Of the *BRAFmut* patients, 5 of 12 received a BRAF inhibitor and mOS in these patients was not reached (CI: NA-NA). mOS in the 7 patients treated without BRAF inhibitor was 28.1 months (CI: 5.5-50.7, p=0.34).

**Analysis of factors driving short survival in *EGFRmut* patients**

To this end we compared patients with below the lower quartile of mOS with the remaining patients (total cohort n=65). Only *TP53* co-mutations, being moderately predictive for shorter survival, were slightly enriched in the lowest quartile, 21.2% of patients had *TP53mut* and 12.5% in the remaining quartiles, but the difference was far from significant (p=0.54). Also there was no association with sex, and PDL-1 TPS (p=0.65, p=0.42).

**Analyzing stability changes**

We used the CUPSAT method for predicting protein stability changes which uses a PDB structure for prediction. As none of the experimental CTNNB1 structures included the crucial residues in exon 3 (e.g. S37), we used the AlphaFold structure of UniProt ID P35222 from the AlphaFold Protein Structure Database. However, the prediction of exon 3 is impaired as shown in S3. Thus, the stability prediction results have to be interpreted with caution.

### Figures

**Supplementary Figure S1**

PDL-1 expression compared across *CTNNB1* mutation positions and color-coded by substituting amino acid. Black bars indicate the median.

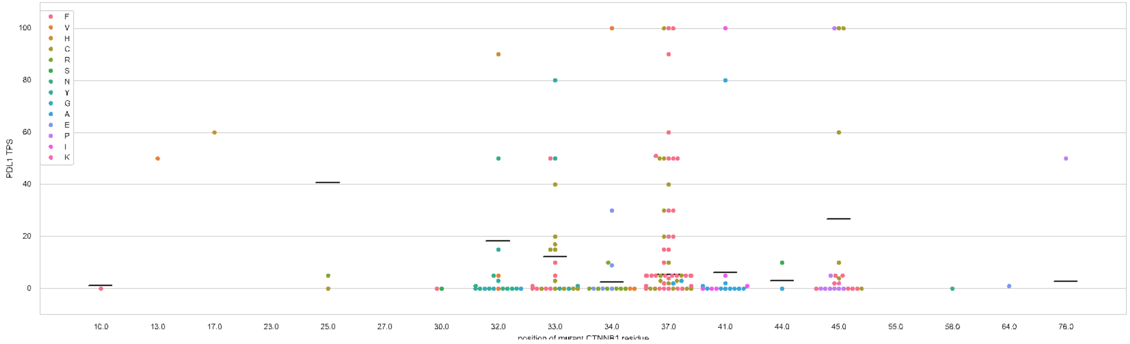

**Supplementary Figure S2**

AlphaMissense score difference between *CTNNB1* resistant vs non resistant mutations.

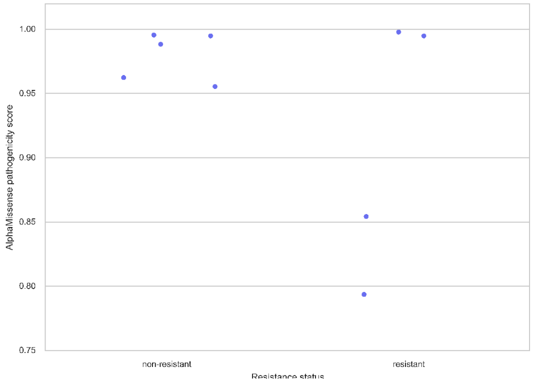

**Supplementary Figure S3**

Note that as AlphaFold2 and AlphaMissense share crucial architecture parts (Evoformer for information extraction from Multiple Sequence Alignments), erroneous AlphaFold2 structures may also lead to impaired AlphaMissense predictions, as is the case with CTNNB1. AlphaFold2 and AlphaFold3 predictions are similarly distorted, and AlphaFold2 was used as the basis for stability prediction.

AlphaFold predicted aligned error:

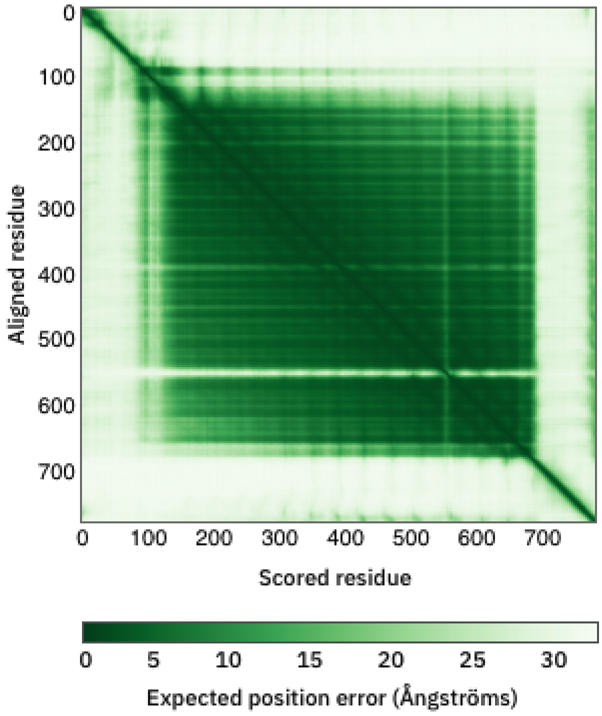

Heatmap visualizing AlphaMissense scores across the entire CTNNB1 protein, showing likewise affected scores:

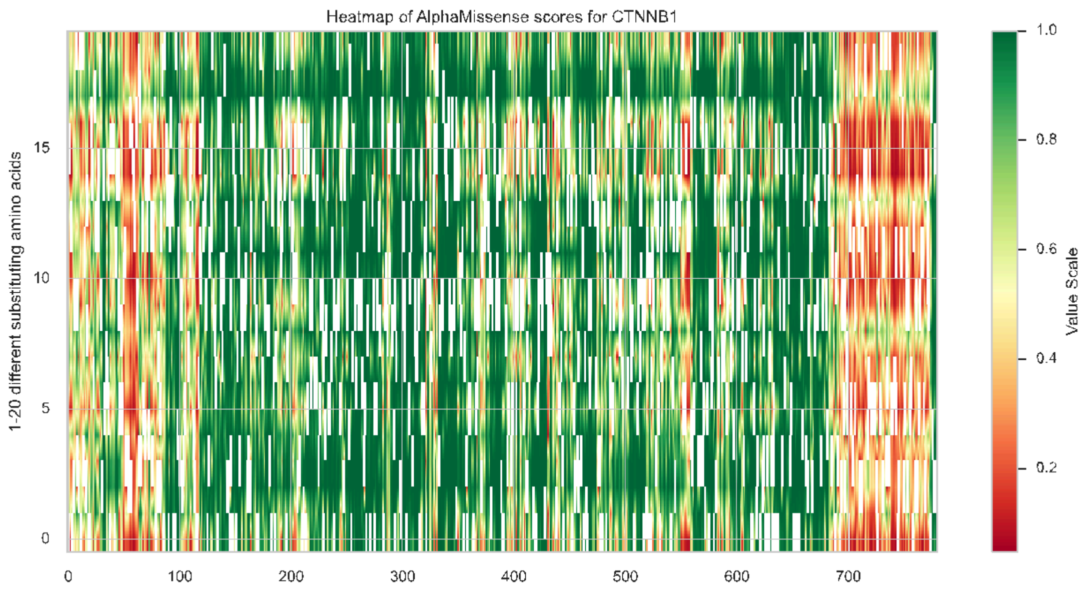

PDB Structure 3ouw color-coded by mean AlphaMissense scores at the respective position (red - pathogenic score of 1, blue - benign score of 0):

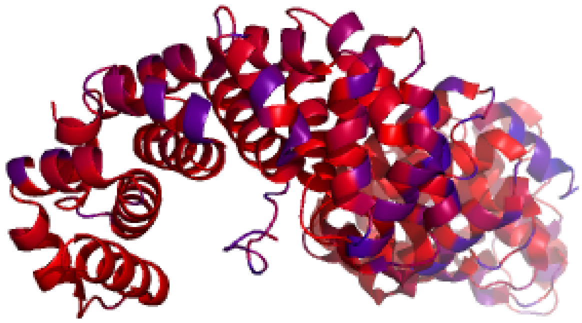

**Supplementary Figure 4**

All of the *PIK3CA* mutations known (n=28/32, 87.5%) to OncoKB were annotated oncogenic and gain-of-function. E545K was the most frequent mutations (53.1%, n=17/32). Lollipop chart of *PIK3CA* mutations:

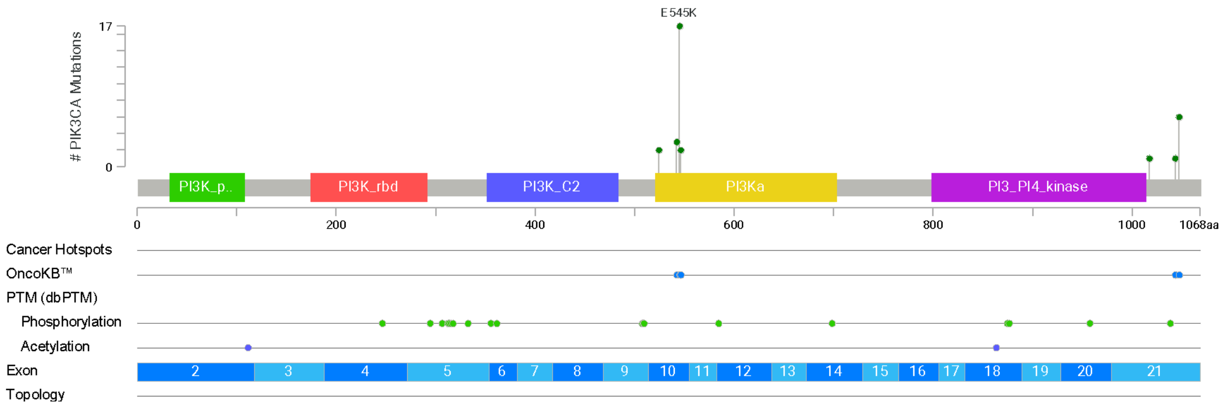

#### Supplementary Figure 5

Absolute counts and clustering of co-mutations which co-occur together with other co-mutations:

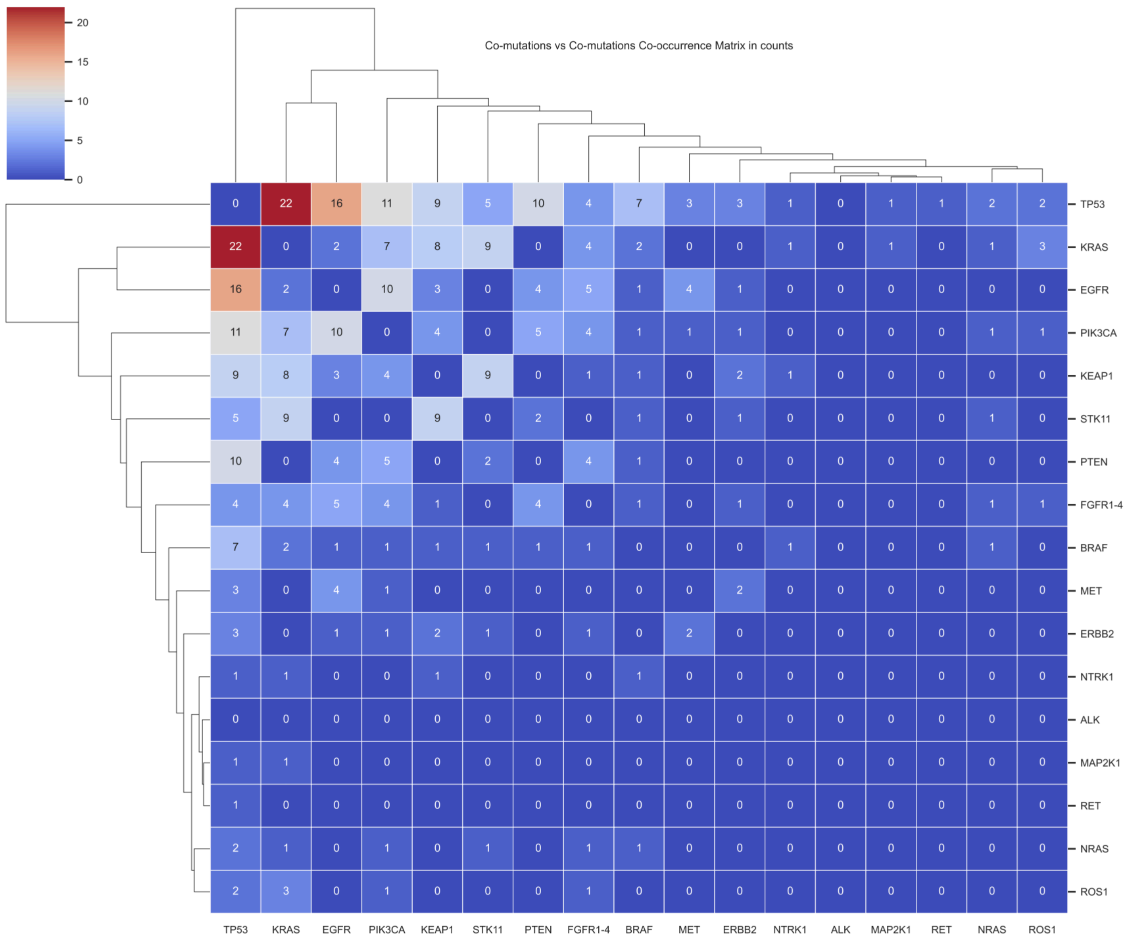
Co-mutations vs Co-mutations Co-occurrence Matrix in counts
20
15
10
5
0
TP53 0 22 16 11 9 5 10 4 7 3 3 1 0 1 1 2 2
KRAS 22 0 2 7 8 9 0 4 2 0 0 1 0 1 0 1 3
EGFR 16 2 0 10 3 0 4 5 1 4 1 0 0 0 0 0 0
PIK3CA 11 7 10 0 4 0 5 4 1 1 1 0 0 0 0 1 1
KEAP1 9 8 3 4 0 9 0 1 1 0 2 1 0 0 0 0 0
STK11 5 9 0 0 9 0 2 0 1 0 1 0 0 0 0 1 0
PTEN 10 0 4 5 0 2 0 4 1 0 0 0 0 0 0 0 0
FGFR1-4 4 4 5 4 1 0 4 0 1 0 1 0 0 0 0 1 1
BRAF 7 2 1 1 1 1 1 1 0 0 0 1 0 0 0 1 0
MET 3 0 4 1 0 0 0 0 0 0 2 0 0 0 0 0 0
ERBB2 3 0 1 1 2 1 0 1 0 2 0 0 0 0 0 0 0
NTRK1 1 1 0 0 1 0 0 0 1 0 0 0 0 0 0 0 0
ALK 0 0 0 0 0 0 0 0 0 0 0 0 0 0 0 0 0
MAP2K1 1 1 0 0 0 0 0 0 0 0 0 0 0 0 0 0 0
RET 1 0 0 0 0 0 0 0 0 0 0 0 0 0 0 0 0
NRAS 2 1 0 1 0 1 0 1 1 0 0 0 0 0 0 0 0
ROS1 2 3 0 1 0 0 0 1 0 0 0 0 0 0 0 0 0
TP53 KRAS EGFR PIK3CA KEAP1 STK11 PTEN FGFR1-4 BRAF MET ERBB2 NTRK1 ALK MAP2K1 RET NRAS ROS1

### Table

Supplementary Table: **NGS LUN Panels**

| **NGS method** | **LUN5 PANEL** | **regions sequenced** | **LUN6 PANEL** | **regions sequenced** |
|---|---|---|---|---|
| genes sequenced (n) | 19 | | 26 | |
| genes | ALK | 22 - 25 | ALK | 21-25 |
| | BRAF | 11. 15 | BRAF | 11. 15 |
| | CTNNB1 | 3 | CTNNB1 | 3 |
| | EGFR | 18 - 21 | EGFR | 18 - 21 |
| | ERBB2 | 8. 19. 20 | ERBB2 | 9. 19. 20 |
| | FGFR1 | 4 - 7. 10. 12 - 15 | FGFR1 | 4-7. 10. 12-15 |
| | FGFR2 | 6 - 15. 18 | FGFR2 | 6-15, 18 |
| | FGFR3 | 3. 7. 9. 10 (codon 429 - 471). 12 (codon 512 - 529). 14. 16. 18 (codon 769 - 807) | FGFR3 | 3, 6, 7, 9, 10, 12, 14, 16, 18 |
| | FGFR4 | 3. 6. 9. 12. 13 | FGFR4 | 3, 6, 9, 12, 13, 15, 16 (Codon 672-712) |
| | IDH1 | 4 | HRAS | 2-4 |
| | IDH2 | 4 | IDH1 | 4 |

| NGS method | LUN5 PANEL | regions sequenced | LUN6 PANEL | regions sequenced |
|---|---|---|---|---|
| | KRAS | 2 - 4 | IDH2 | 4 |
| | MAP2K1 | 2. 3 | KEAP1 | 2 - 6 |
| | MET | 14. 16 - 19 | KRAS | 2 - 4 |
| | NRAS | 2 - 4 | MAP2K1 | 2. 3 |
| | PIK3CA | 10. 21 | MET | 14. 16 - 19 |
| | PTEN | 1 - 8 | NRAS | 2 - 4 |
| | ROS1 | 34 - 41 | NTRK1 | 13-17 |
| | TP53 | 4 (codon 97 - 125). 5. 6. 7. 8 | NTRK2 | 16-19 |
| | | | NTRK3 | 15-20 |
| | | | PIK3CA | 8. 10. 21 |
| | | | PTEN | 1 - 8 |
| | | | RET | 10-18 |
| | | | ROS1 | 34-41 |
| | | | STK11 | 1-9 |
| | | | TP53 | 2-11 |
